# School Feeding Programmes and Physical Nutrition Outcomes of Primary School Children in Developing Countries

**DOI:** 10.1101/2022.04.19.22274039

**Authors:** Mustapha Titi Yussif, Vincent Awuah Adocta, Charles Apprey, Reginald Adjetey Annan, Prosper Galseku

## Abstract

**Context:** School feeding programmes have been widely implemented and particularly in developing countries with the aim to improve school enrolment and attendance especially of girls and to reduce short term hunger to improve children’s performance in school.

Beyond the first 1000 days of the lives of children, school feeding programmes remain one of the critical interventions that have used schools as a platform to contribute to the fulfilment of their nutritional needs though the evidence to this effect is little and mixed.

**Objective:** This review focused on assessing the impact of school feeding programmes on reduction in underweight, thinness, and stunting among primary school children in developing countries.

**Data sources:** Electronic searches were carried out in PUBMED, SCORPUS and Cochrane library. The WHO clinical trials registry as well as reference lists of relevant articles were also hand searched.

**Data Extraction:** Data was extracted from included studies which have been published in the past 10 years (2010 – August 2021) from original research where the main intervention was the provision of school based meals.

**Data analysis:** Meta-analysis was conducted to determine changes in height-for-age (HAZ), weight-for-age (WAZ) and BMI-for-age (BAZ) z scores. A random effects model was applied to determine the mean difference in all outcomes of interest which were evaluated as continuous variables.

**Results:** Children aged 3 – 16 years were enrolled in the included studies and the number of participants ranged between 321 and 2,869 across studies. Of the included studies, the feeding intervention provided for a minimum of 30% RDA for the age group with the intervention lasting up to a maximum of 34 weeks. The impact of school feeding intervention on HAZ, BAZ and WAZ showed statistically non-significant (p>0.05) mean differences of 0.02 (95% CI, -0.06 to 0.10), 0.11 (95% CI, -0.01 to 0.23) and 0.06 (95% CI, -0.04 to 0.16) respectively

**Conclusion:** School feeding interventions have not shown any significant positive effect on the physical nutrition outcomes of primary school children. Short duration of intervention of studies, poor compliance to feeding and substitution of school meals could have accounted for the weak effect sizes.

## INTRODUCTION

### Background

Poor diet is the leading cause of mortality and morbidity worldwide, exceeding the burdens attributable to many other major global health challenges. According to the 2020 Global Nutrition Report^1^, an average of 820 million people worldwide (1 in every 9) are hungry or undernourished, with numbers rising since 2015, especially in Africa, West Asia and Latin America.

This situation of poor nutrition and health conditions in these developing regions apart from the most often fatal end, have implications for the later lives of the surviving children. It is known that malnutrition worsens poverty and reduces economic growth and its said to cost many developing nations 2-3% of their GDP every year^2^.

Malnutrition during early childhood results in children achieving less at school during school age and their productivity and state of health in adult life becomes seriously affected.

Educational and health outcomes however can still be improved for children during their school age by using the school as an opportune setting to provide adequate health and nutrition interventions and services.

School feeding programmes are one of such interventions that have used schools as a platform to contribute to the fulfilment of the nutritional needs of children beyond the first 2 years of their lives. School feeding programmes are thus interventions that provide food to school children either as in-school meals where the children are fed at school or given take-home food rations^3^.

School feeding programmes come in different modalities and are currently being used in over 161 countries providing school meals to over 388 million children^4^ and receiving various forms and magnitude of support from the World Food Programme and its development partners. According to the World Food Programme^4^, the number of children receiving school meals grew by 9 percent globally and 36 percent in low-income countries between 2013 and 2020.

These feeding programmes have been implemented in many developing countries with varying objectives such as the reduction of short term hunger to improve children’s performance in school, encourage school enrolment and attendance, retention of girls in school^5^ and also as a means to improve nutritional status of school aged children^6^.

### Objectives of the review

It is estimated that governments invest between US$41 - US$43 billion annually on school feeding programs^4^ yet there is a huge controversy over the effectiveness of the school feeding programmes (SFPs). Evidence available show remarkable impact of the SFPs in improving school attendance however evidence for nutritional benefits is little and mixed. A recent meta-analysis^7^ shows impact of feeding programmes in schools on iron deficiency anaemia reduction in developing countries.

Following the Preferred Reporting Items for Systematic Reviews and Meta-Analyses (PRISMA) guidelines, this systematic review therefore considered the most recent literature for new evidence from randomized controlled trials on the impact of the school feeding programmes on physical nutritional outcomes of primary school children in developing countries. The review primarily focused on the intervention’s impact on reduction in underweight, thinness, and stunting among primary school children in developing countries.

## METHODS

### Eligibility Criteria

#### Type of Studies

This review included both experimental and observational studies. The randomized controlled trials that met the eligibility criteria were solely used in the quantitative data synthesis in the meta-analysis whilst the observational studies were used together with the trials for the qualitative synthesis.

Only studies written in English and peer reviewed with full text available were included. Studies with only abstract available and those that were not fully written in English language were excluded.

In terms of publication timeframe, this review only included studies which have been published within the past 10 years (from 2010 to August 2021). This was to ensure the review gathered the most current literature in the study area considering new evidence and also not to merely repeat the findings of earlier systematic reviews (Kristjannson et al.^8^ and Lamis et al.^9^) that had a similar focus.

The review also excluded articles which were written based on review of existing literature (systematic reviews and meta-analysis) and thus only included articles based on original research (primary data). Only original research articles have been included as a measure of avoiding duplicate effect of an article associated with its use as primary literature and also its use in a secondary literature in the same work.

#### Type of participants

This review aims to draw conclusions on the impact of school feeding programmes on primary school children in developing countries hence only studies that were conducted in developing countries were included. Developing countries were determined using the World Bank’s list of economies^10^.

The inclusion and exclusion criteria did not segregate between whether the study sample was drawn from an urban, rural, slum or socio-economically disadvantaged areas in as far as the setting is in a low and middle income country.

Studies which were carried out on primary school children were included whiles those that only involved pre-school or high school children were excluded. Inclusion was not narrowed down to any article with sample population of a specific age group but broadly on primary school pupils irrespective of their age or stage.

#### Types of Intervention

Studies were only included if the main intervention was the provision of school based meals through a school feeding programme which includes breakfast or lunch and also snacks including milk and cookies (biscuits) as defined by the World Food Programme^3^.

In studies where more than one interventions was provided, such studies were included in this review and the results of the intervention that fits the definition of school feeding described above was included in the pooled effect analysis. However if all interventions are school based meals, then the outcomes of all the interventions are included in the analysis.

Studies were excluded if the intervention was not in a school setting. Studies that provided only micronutrient supplementation, take-home food rations, food stamps and stand-alone school nutrition education were excluded as well. Studies with interventions which provide cash transfers to parents of primary school children were also excluded from this review.

#### Types of Outcome Measures

Studies that measured changes in physical nutrition outcomes associated with in-school meals were included in this review. Outcomes of physical nutritional status that were considered in this review were the anthropometric indicators of leanness, chronic and acute malnutrition particularly, underweight, stunting and BMI for age.

The Participants, Interventions, Comparators, Outcomes and Study design (PICOS) criteria for inclusion and exclusion are described in Table 1.

**Table 1:** PICOS criteria for inclusion and exclusion of studies.

| PICOS Criteria | Inclusion | Exclusion |
| --- | --- | --- |
| **Participants** | - Study subjects being primary school children<br>- Study participants being from a developing country | - Pre-school children and high school children<br>- Study participants being from a developed country |
| **Interventions** | - School feeding – in-school meals provided; breakfast or lunch or snacks | - Take home food rations<br>- School based micronutrient supplementation<br>- Food stamps |
| **Comparators** | - Articles with an intervention and comparison group/control group<br>- Control/comparison group did receive any kind of | - Control group receiving any form of in-school feeding/micronutrient supplementation |
| **Outcomes** | - Primary: studies that measured indicators of iron deficiency anaemia (haemoglobin and serum ferritin concentrations)<br>- Secondary: dietary iron consumption from in-school meals |  |
| **Study Design** | - Randomized trials<br>- Observational studies; to be used for only qualitative synthesis | - Conference abstracts of randomized trials without sufficient information |

#### Sources of Information

The studies included were identified through the use of electronic database search. The databases that were particularly searched were COCHRANE LIBRARY, PUBMED and SCORPUS.

In order to explore grey literature, the World Health Organization (WHO) international clinical trials registry platform (http://apps.who.int/trialsearch/) was also searched for studies which have been completed but not published in peer review journals.

#### Search Strategy

The basic search terms that were used were, ‘school lunch’, ‘school feeding’, ‘mid-day meal’, ‘school based feeding’, ‘school nutrition’. Some additional terms that were used included ‘nutritional status’, ‘anthropometry’, ‘stunting’, wasting’, ‘underweight’. The basic search terms were either used individually in the search or in combination with the additional search terms to generate the articles. Table 2 shows the detailed search strategy as applied in COCHRANE LIBRARY, PUBMED and SCORPUS.

**Table 2:**
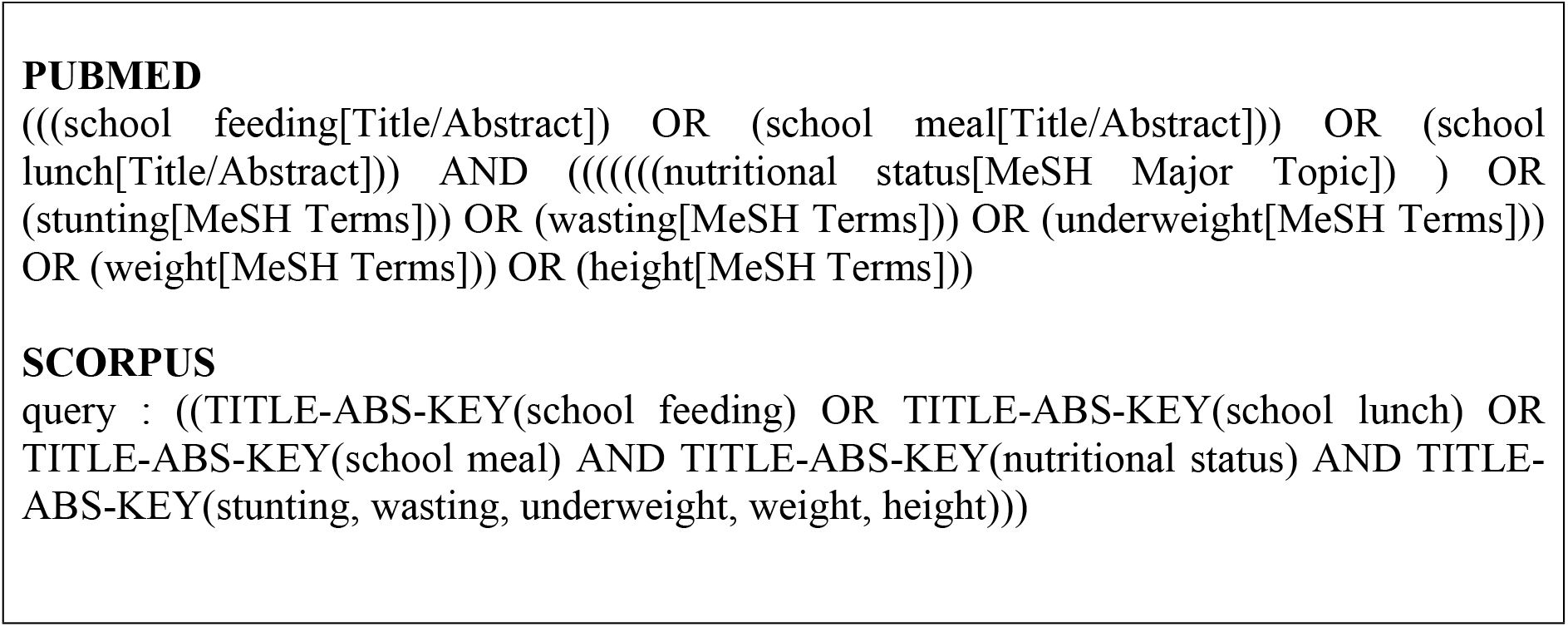
Search Strategy.

### Data Collection Process

Data collection forms were developed a priori using the Covidence software and in a way that allowed eligibility to be assessed and that extracted data responded to the research questions. There was a pilot-testing of the data extraction forms using two included articles. This was to ensure that the data collection forms generated were easy to use and that data extracted was consistent, clear and complete. Eligibility assessment was done by two persons independently and disagreements resolved by consensus.

Data was extracted on study characteristics such as year, country of origin, study design, sample sizes for intervention and control groups, gender, age and length of intervention. Description of the intervention and outcome measurement as well as key findings and data on direction of results were also extracted

### Selection of studies and data synthesis

The results generated from the use of the search terms was filtered using the various inclusion and exclusion criteria described in Table 1 above in each of the electronic databases.

The total number of articles retrieved after preliminary search from the three databases were 493. 2 other articles were obtained from the search for grey literature. Upon the removal of 54 duplicate articles, 413 irrelevant articles were further removed through tittle and abstract screening.

Full text reading was done for 28 studies during which an additional 20 articles were excluded with 8 articles remaining which satisfied the inclusion criteria. Of the 8 included studies, 4 were randomised controlled trials whiles the other 4 were observational studies. Figure 1 details the selection of studies.

**Figure 1:**
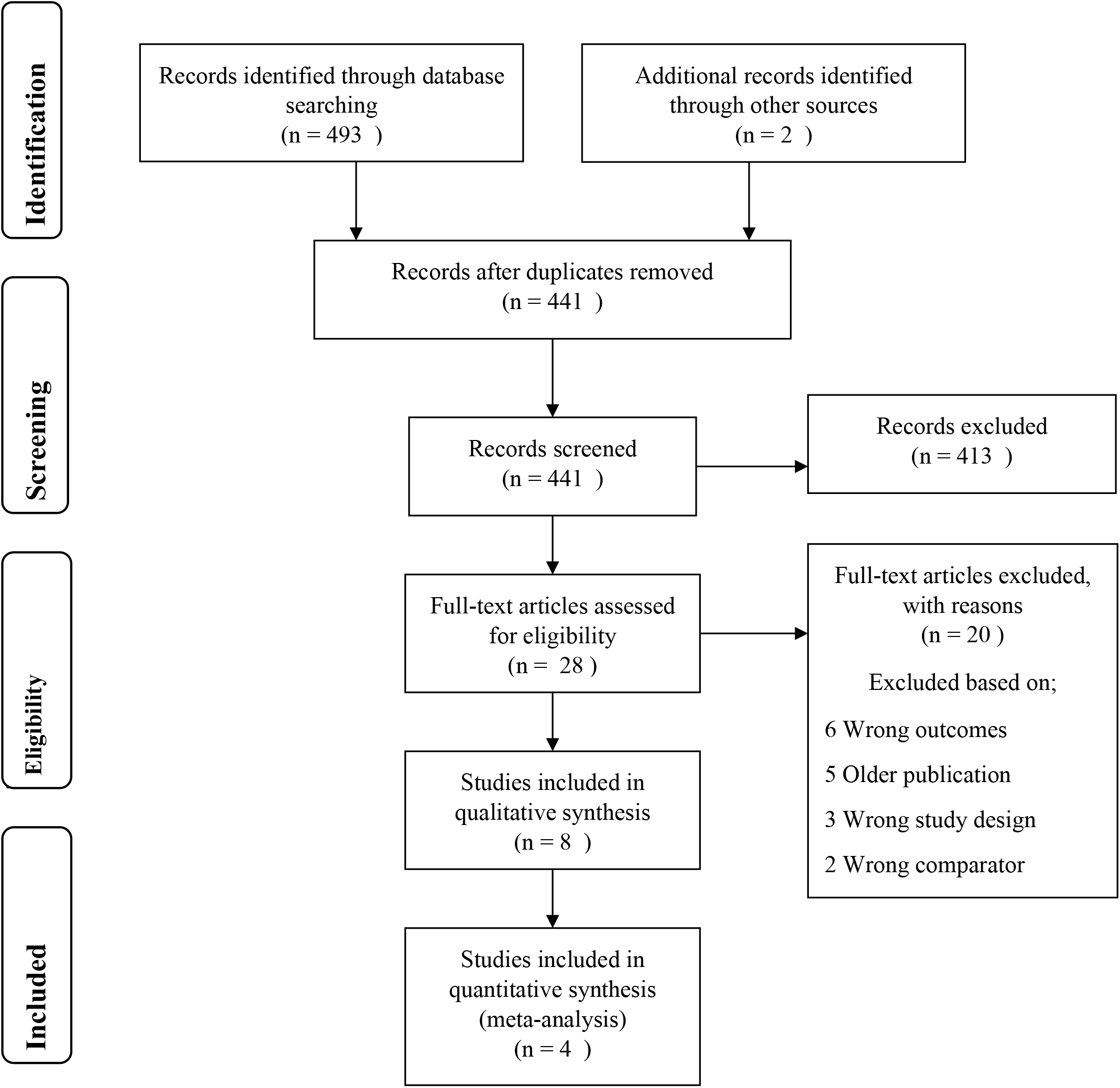
Flow Diagram of Literature Search Process.

### Summary measures and synthesis of results

Meta-analysis was conducted using Cochrane Collaboration Review Manager (RevMan version 5.4). Predicting an eminent clinical heterogeneity in the studies related to the varying modes of the school feeding intervention, a random effects model was applied in the analysis and this was determined a priori.

All the three outcomes of this review; Height-for-age z scores and Weight-for-age z scores as well as BMI-for-age z scores were evaluated as continuous outcomes using mean differences. In one of the studies; Neervort et al.^11^, all the outcome data was presented in prevalence rates and were thus not used in the meta analysis because the mean and standard deviations could not be obtained.

For studies that provided standard errors instead of standard deviations, the standard deviations for the sample means were calculated using SD = SE x √N where SD is Standard Deviation, SE, Standard error and N, the sample size^12^.

### Assessing Heterogeneity and Publication Bias

Clinical heterogeneity was assessed by considering the ages of the participants enrolled in the study and the type and quality of meals served in the school feeding programmes used in the various studies. Statistical heterogeneity on the other hand was assessed using visual inspection of forest plots, I^2^ measures and their respective chi square tests^13^. An I^2^ greater than 75% was evaluated as considerable heterogeneity^14^.

Funnel plots were not considered in the assessment of publication bias due to the few number of studies used in the meta analysis^15^. However, publication bias was reduced through the adoption of a robust search strategy which included a thorough hand search of references of included studies as well as searches on the websites of the World Food Programme and the WHO clinical trials registry to identify studies that were not published in peer review journals.

## RESULTS

### Characteristics of included studies

The review included studies that were conducted in developing countries across three continents; Africa, Asia and Latin America. Five of the studies were conducted in Africa, two studies in Latin America and One study in Asia.

Participants enrolled in the studies that were included in the review were all primary school pupils aged between 3 and 16 years with number of participants ranging from 321 to 2,869 across studies. With respect to the number of participants used in the meta analysis to evaluate the impact of school feeding programmes on the various physical nutritional outcomes, 3955 participants included in the trials was used to evaluate programme impact on height-for-age z scores (stunting) whilst 1562 and 3930 included in the trials were used to evaluate impact on weight-for-age and BMI-for-age respectively as shown in Table 3.

**Table 3:** Effect estimates on outcomes.

| Outcome or Subgroup | Studies | Participants | Statistical Method | Estimate Effect |
| --- | --- | --- | --- | --- |
| **1.1 Height-for-age z scores** | 4 | 3955 | Mean Difference (IV, Random, 95% CI) | 0.02 [-0.06, 0.10] |
| **1.2 BMI-for-age z scores** | 4 | 3930 | Mean Difference (IV, Random, 95% CI) | 0.11 [-0.01, 0.23] |
| **1.3 Weight-for-age z scores** | 3 | 1562 | Mean Difference (IV, Random, 95% CI) | 0.06 [-0.04, 0.16] |

The trials that were included in this review administered school feeding interventions that provided either snacks or a hot meal for lunch made from local foods. The feeding interventions provided for a minimum of 30% of the Recommended Dietary Allowance for the age group of the children enrolled with the interventions lasting for a minimum of 13 weeks to a maximum of 34 weeks.

### Risk of Bias in Included Studies

For the randomized control trials which have been included in this review, the risk of bias judgements have been shown in the ‘risk of bias’ summary tables and ‘risk of bias’ graph in Figure 2 and Figure 3 respectively.

**Figure 2:**
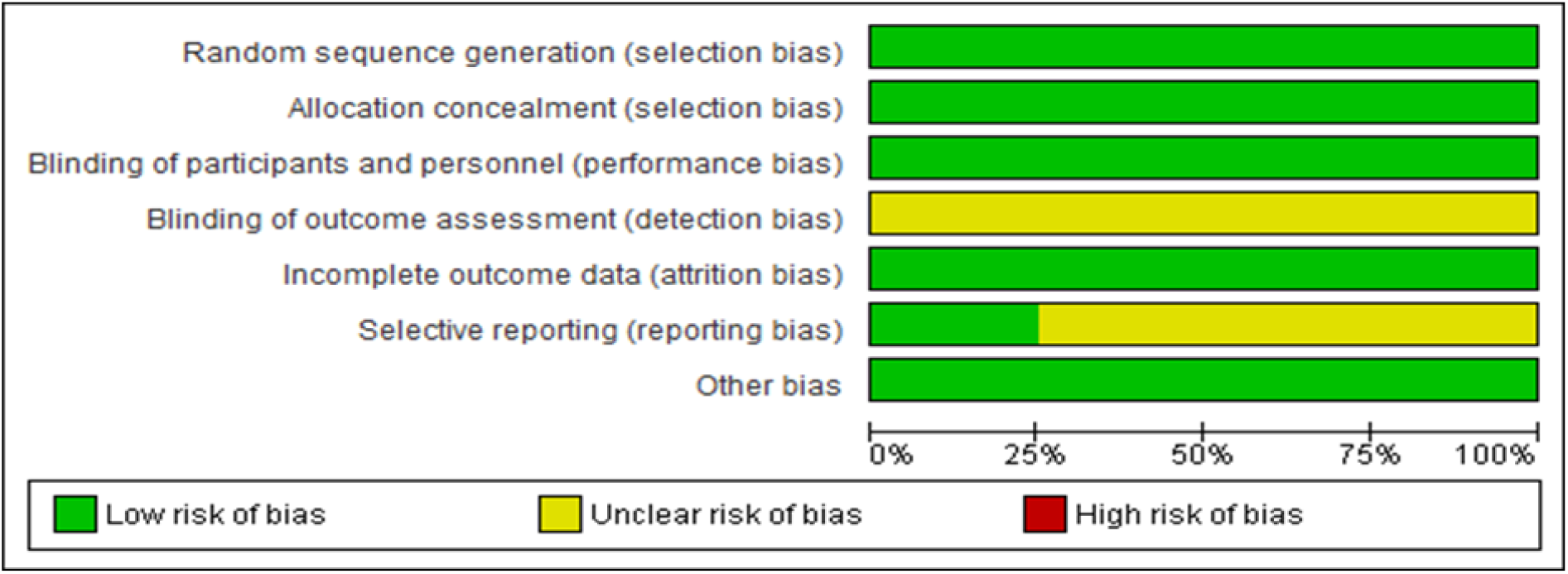
Risk of bias graph: review authors’ judgements about each risk of bias item presented as percentages across all included studies.

**Figure 3:**
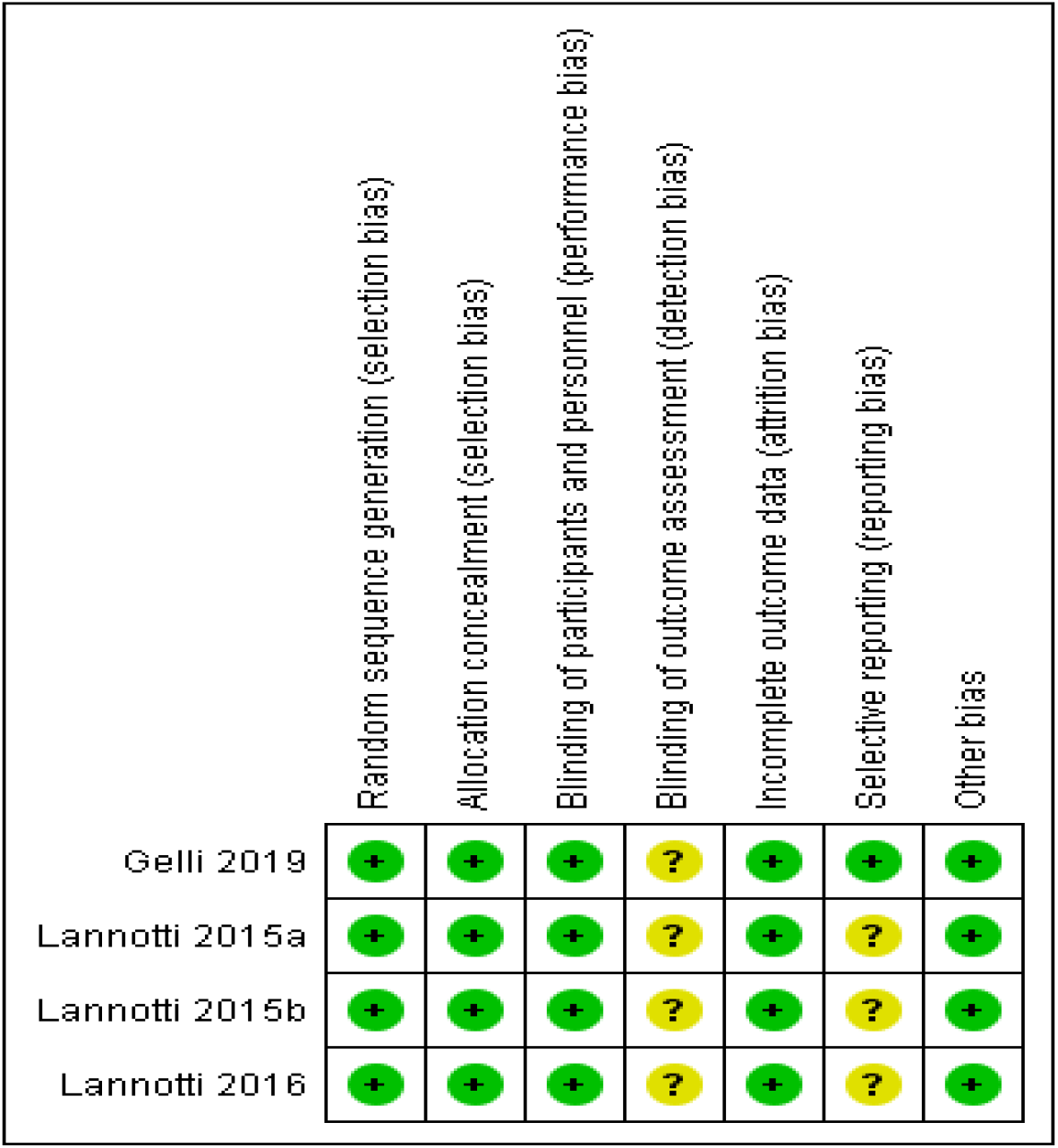
Risk of bias summary: review authors’ judgements about each risk of bias item for each included study.

**Figure 4:**
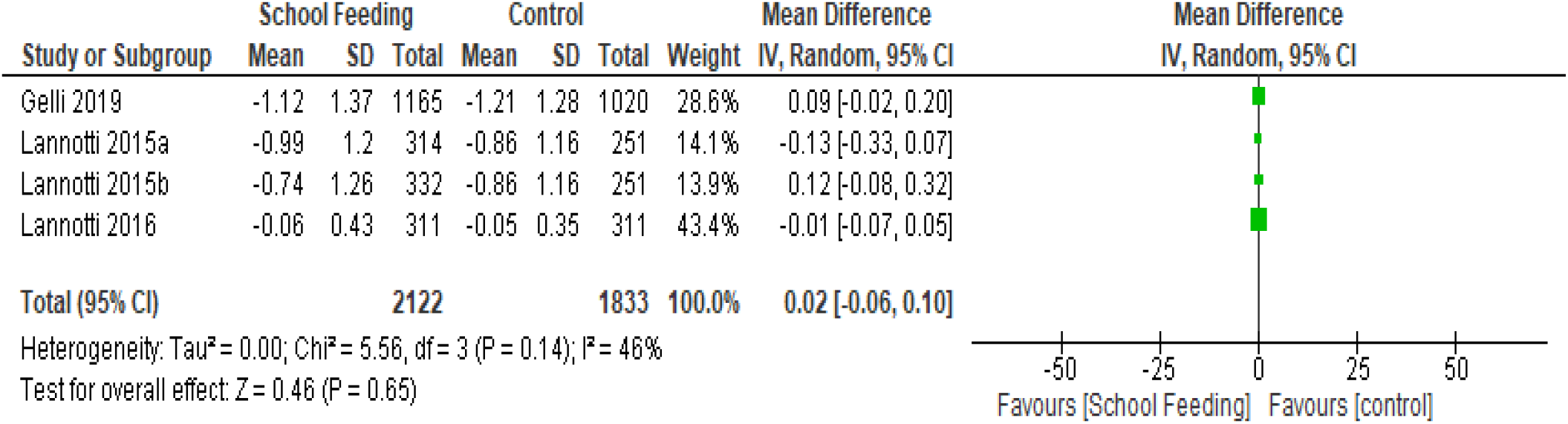
Forest plot of school feeding vrs control, RCTs, Outcome: Height-for-age z scores (HAZ)

**Figure 5:**
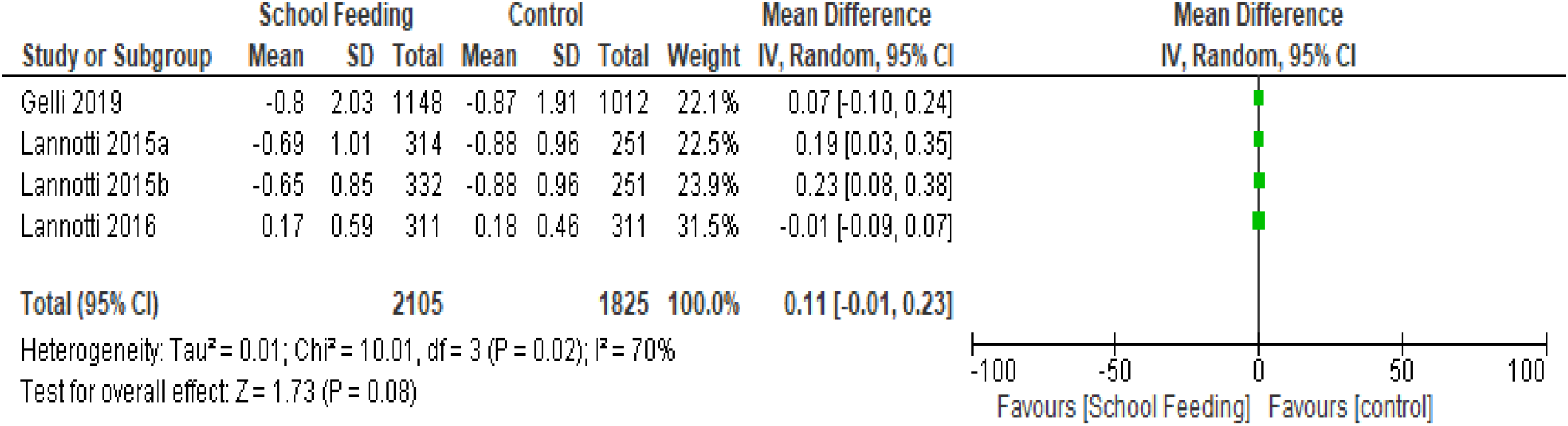
Forest plot of school feeding vrs control, RCTs, Outcome: BMI-for-age z scores (BAZ)

**Figure 6:**
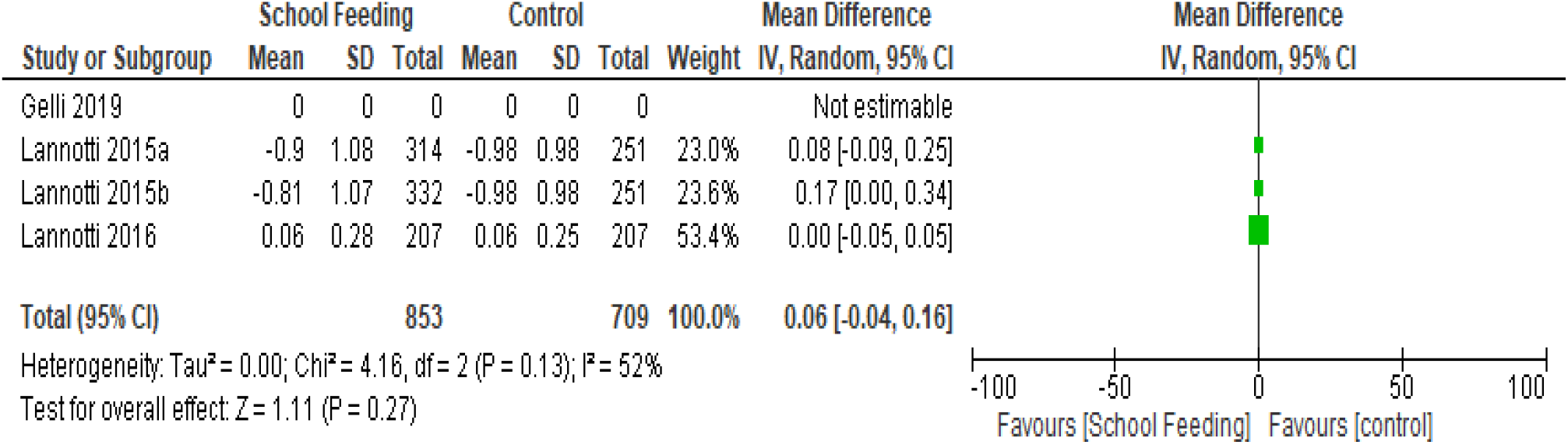
Forest plot of school feeding vrs control, RCTs, Outcome: Weight-for-age z scores (WAZ)

All the included trials described random sequence generation and allocation concealment adequately and thus were rated as low risk of bias.

Blinding of participants and personnel was only described by Gelli et al.^16^. It was stated that participants and survey enumerators were not blinded to the allocation whiles the remaining studies were silent on this. Not this withstanding, all the studies were judged to be of low risk of bias for blinding of participants because the outcomes of interest are not likely to be influenced by a lack of blinding.

For blinding of outcome assessment, all the studies were judged to have an unclear risk of bias for blinding because none of the studies had reported on it.

All the trials included in this review were judged to have a low risk of bias associated with incomplete outcome data or attrition bias.

Only Gelli et al.^16^ had published the protocol for the study which was then used to confirm that the outcomes that were reported were same as was published in the protocol. Thus a low risk of bias was noted for the study conducted by Gelli et al.^16^ whiles the remaining three trials^17-19^ were judged to have an unclear risk of risk of reporting bias as their protocols could not be retrieved hence there was no enough information available to serve as basis of judgment.

On other bias, preference bias was the other potential sources of bias that was evaluated. All the studies were rated as having a low risk of other bias.

### Synthesis of School Feeding Programme Impact

#### Height-for-Age z-score

In order to evaluate the impact of school feeding programmes on height-for-age z scores as a marker for stunting, 3 randomized controlled trials with a total of 3,955 participants were included in the Meta analysis. One of the studies; Lannotti et al.^17^, provided a two arm intervention and both arms were used separately in the analysis. 3 of the interventions from the trials that were included in this analysis provided snacks while 1 trial provided a hot meal lunch. This indicates that the studies that contributed to the pooled effect were very similar and as well not statistically heterogeneous (Tau2 = 0.00, chi2 = 5.56, df= 3, p =0.14; I^2^ = 46%) as indicated in the meta analysis.

The result of the meta analysis indicates that there is no significant effect (p = 0.65) of school feeding programmes on height-for-age z-scores (HAZ) as the intervention only resulted in a marginal effect size of 0.02 (95% CI, -0.06 to 0.10) in HAZ among school feeding beneficiaries compared to non-beneficiaries.

#### BMI-for-Age z-score

Two single arm and one double arm randomized control trials were included in the meta analysis to evaluate the impact of school feeding programmes on BMI-for-Age z-scores among primary school children in developing countries. A total of 3930 participants were involved with feeding interventions lasting for an average of 24 weeks where either energy dense snacks or a hot meal lunch made from local ingredients were used.

From the meta analysis, school feeding does not seem to confer any significant impact in terms of BAZ on beneficiaries. An insignificant (p = 0.08) mean difference of 0.11(95% CI, - to 0.23) BAZ exists between the school feeding group and the controls. The contribution of the various trials to the net effect was fairly homogenous at a Tau2 of 0.01, chi2 = 10.01 with df= 3, p =0.02; I^2^ = 70% requiring no sub group analysis.

#### Weight-for-age z-scores

A total of 1562 participants were included from two trials in the Meta analysis to determine the effect of providing school feeding programmes to primary school children on their weight for age z scores (WAZ). The two trials of which one had a two arm intervention were both conducted in Latin America where the intervention groups were given snacks that provided 165 kcal and 260kcal over an average period of 19 weeks.

The Meta analysis which indicated that heterogeneity among trials that contributed to the pooled effect was at Tau2 of 0.00, chi2 = 4.16 with df= 2, p =0.13; I^2^ = 52% provided no substantial evidence to conclude that the provision of snacks in a school feeding programme to primary school children will improve their WAZ. The children that were provided with the snacks only had a WAZ of 0.06 (95% CI, -0.04 to 0.16) more than their counterparts who did not receive the snacks. This difference has been shown to be statistically insignificant (p = 0.27).

## DISCUSSION

Full text screening of 28 articles resulted in the inclusion of 8 studies in this review of which 4 studies were randomized controlled trials which were used in the Meta analysis leading to the main conclusions in this review. The studies were conducted in eight developing countries across Africa, Asia and Latin America where the feeding interventions provided in the trials were either snacks or lunch made from local recipes.

The meta-analysis of three homogenous randomized controlled trials which included 3,955 primary school children showed that the provision of school feeding programmes does not result in any significant change in height-for-age z-scores and thus does not translate to a reduction in stunting among beneficiaries vis a vis non-beneficiaries.

The four observational studies that were included in this review all evaluated programme impact on height-for-age z scores however with mixed results. Two large studies;Wang et al.^20^ and Abizari et al.^21^, conducted in China and Ghana respectively found that school feeding programmes did not influence any significant change in HAZ and stunting prevalence. In contrast, a study conducted by Kwabla et al.^22^ in Ghana found prevalence rates of stunting to be lower (16.2%) among school feeding beneficiaries than non-programme participants (17.2%). Another study conducted in Ethiopia; Zenebe et al.^23^, found HAZ to be significantly higher (p<0.001) among school feeding participants with a mean difference of 0.57 (95% CI: 0.36 – 0.78).

BMI-for-age z scores is a measure of thinness among children. This review has shown that for primary school children, the provision of meals/snacks in school does not confer any positive impact on their thinness status. The Meta analysis that combined three trials that provided snacks and hot meal lunches to 3930 children in school over an average of 24 weeks period did not find any significant impact on BAZ.

A similar conclusion can be arrived at when the observational studies included in this review are considered. Three out of four observational studies found no significant positive influence of school feeding programmes on BAZ. In fact, one of the three studies; Kwabla et al.^22^, found that the prevalence of thinness was 2 times higher (9.3%) among children in schools providing school feeding programmes compared to the non-school feeding programme schools (4.6%).

Weight-for-age z-scores (WAZ) is one of the commonest indicators of nutritional status among children. It classifies children either as underweight or not and is a composite indicator of co-existence of both chronic and acute malnutrition. From the Meta analysis conducted in this review using two trials with three intervention arms that provided snacks on daily basis to school children for an average period of 19 weeks, the interventions were found not to significantly increase WAZ among beneficiaries. Thus, school feeding programmes that provide snacks to children does not significantly impact on underweight status of children in primary schools.

The only observational study in this review that evaluated the impact of school feeding programmes on WAZ; Abizari et al.^21^,found no significant difference in prevalence of underweight between children benefiting from the feeding programme and non-beneficiaries thus affirming the conclusion arrived by the results of the meta analysis.

## CONCLUSION

This review has demonstrated through the results of the meta analysis of four randomized controlled trials that school feeding programmes do not have any significant impact on physical nutrition outcomes particularly height-for-age z scores, BMI-for-age z scores and weight-for-age z scores. Evidence from observational studies generally support the conclusions of the meta analysis with a little mixed evidence of the impact of school feeding programmes on stunting reduction in primary school children.

Although, the trials that were included in the review were mostly similar and the Meta analysis results depicted considerable statistical homogeneity in all cases, the average period of the feeding intervention of 27 weeks may be short and might have resulted in the insignificant effect sizes.

Also, most of the studies included in the review did not report on how compliance was monitored or enforced thus, a possible low compliance to the feeding interventions could have also compromised programme impacts.

Another factor that could affect programme impact is substitution; where the home diet may be reduced for children who are benefiting from school feeding. Though none of the studies in this review provided data on this, other studies have reported it among poor families^24,25^.

Studies that aim to evaluate the impact of school feeding programmes on the nutritional wellbeing of children are of great importance in public health as poor nutrition and health at school age is common especially in developing countries and given that malnutrition is an intergenerational issue. Such studies are equally vital to inform national governments, global food and nutrition stakeholders who invest in these programmes on its effectiveness.

There is therefore the need for more randomized controlled trials which are of high methodological quality to be able to attribute causality and to be eligible to be used in a larger meta-analysis to confirm or otherwise reject the evidence produced by this review.

## Data Availability

Additional data can be obtained from the corresponding author

## ACKNOWLEDGEMENTS

We wish to acknowledge the support received from the Covidence Team who provided a waiver of user fees associated with the use of the Covidence Software which was largely used to manage the systematic review. RevMan, a software from the Cochrane Collaboration was also used for the quantitative synthesis of the review data.

## Funding and Sponsorship

No funding of any sort was received from any third parties or organizations.

## Declaration of Interest

The authors have no relevant interest to declare.

## LIST OF SUPPORTING INFORMATION

1. Summary Table of Included Studies – Randomized Controlled Trials
2. Summary Table of Included Studies – Observational Studies

## Notes

### Competing Interest Statement

The authors have declared no competing interest.

### Funding Statement

No funding was obtained for this study

## REFERENCES

1. Micha, Renata, Mannar, Venkatesh, Afshin, Ashkan, Allemandi, Lorena, Baker, Phillip, Battersby, Jane, et al., 2020 Global nutrition report: action on equity to end malnutrition. Technical Report. Behrman, Nina, ed. Development Initiatives, Bristol, UK. 2020; ISBN 978-1-9164452-7-7. [Monograph] Official URL: https://globalnutritionreport.org/reports/2020-glo…

2. Horton, Susan, Meera Shekar,Christine McDonald, Ajay Mahal and Jana Krystene Brooks. Scaling Up Nutrition: What Will It Cost? World Bank: Washington, DC. 2010

3. World Food Programme. State of School Feeding Worldwide. World Food Programme; 2013: Italy

4. World Food Programme. State of School Feeding Worldwide. World Food Programme;Via C.G. Viola, 2020; 68–70, Rome 00148, Italy

5. Hall A, Hanh TT, Farley K, Quynh TP, Valdivia F. An evaluation of the impact of a school nutrition programme in Vietnam. Public Health Nutr. 2007 Aug;10(8):819–26. doi: 10.1017/S1368980007382530. Epub 2007 Mar 8. PMID: 17381906.

6. Adelman, Sarah & Gilligan, Daniel & Lehrer, Kim. The Impact of Alternative Food for Education Programs on Learning Achievement and Cognitive Development in Northern Uganda. 2008

7. Yussif MT, Vong L, Pilkington K. The Impact of School Feeding Programmes in Reducing Iron Deficiency Anaemia among Primary School Children in Developing Countries: A systematic Review and Meta-Analysis of Randomized Controlled Trials. J Hum Nutr. 2020; 4(1): 87 – 105

8. Kristjansson EA, Robinson V, Petticrew M, MacDonald B, Krasevec J, Janzen L, Greenhalgh T, Wells G, MacGowan J, Farmer A, Shea BJ, Mayhew A, Tugwell P. School feeding for improving the physical and psychosocial health of disadvantaged elementary school children. Cochrane Database Syst Rev. 2007; 1:CD004676.

9. Jomaa LH, McDonnell E, Probart C. School feeding programs in developing countries: impacts on children’s health and educational outcomes. Nutr Rev. 2011; 69(2):83–98.

10. World Bank. World Bank List of Economies 2021.World Bank 2021.Available at: https://datahelpdesk.worldbank.org/knowledgebase/articles/906519-world-bank-country-and-lending-groups accessed on17-08-2021

11. Neervoort F, von Rosenstiel I, Bongers K, Demetriades M, Shacola M, Wolffe rs I. Effect of a school feeding programme on nutritional status and anaemia in an urban slum: a preliminary evaluation in Kenya. J Trop Pediatr.2013; 59 (3):165–74.

12. Hozo SP, Djulbegovic B, Hozo I. Estimating the mean and variance from the median, range, and the size of a sample. BMC Medical Research Methodology. 2005; 5: 13.

13. Higgins JP, Thompson SG, Deeks JJ, Altman DG. Measuring inconsistency in meta-analyses. BMJ. 2003 Sep 6;327(7414):557–60. doi: 10.1136/bmj.327.7414.557. PMID: 12958120; PMCID: PMC192859.

14. Higgins JM, Thompson SG. Quantifying heterogeneity in a meta-analysis. 2002; 21: 1539–1558.

15. Egger M, Davey Smith G, Schneider M, et al. Bias in meta-analysis detected by a simple, graphical test. BMJ 1997; 315: 629–634.

16. Gelli, A., Aurino, E., Folson, G., Arhinful, D., Adamba, C., Osei-Akoto, I., … Alderman, H. A school meals program implemented at scale in Ghana increases height-for-age during midchildhood in girls and in children from poor households: A cluster randomized trial. Journal of Nutrition. 2019; 149(8): 1434–1442. https://doi.org/10.1093/jn/nxz079

17. Iannotti LL, Henretty NM, Delnatus JR, Previl W, Stehl T, Vorkoper S, Bodden J, Maust A, Smidt R, Nash ML, Tamimie CA, Owen BC, Wolff PB. Ready-to-use supplementary food increases fat mass and BMI in Haitian school-aged children. J Nutr. 2015 Apr;145(4):813–22. doi: 10.3945/jn.114.203182. Epub 2015 Feb 11. PMID: 25833784

18. Iannotti LL, Henretty NM, Delnatus JR, Previl W, Stehl T, Vorkoper S, Bodden J, Maust A, Smidt R, Nash ML, Tamimie CA, Owen BC, Wolff PB. Ready-to-use supplementary food increases fat mass and BMI in Haitian school-aged children. J Nutr. 2015 Apr;145(4):813–22. doi: 10.3945/jn.114.203182. Epub 2015 Feb 11. PMID: 25833784

19. Lannotti, L., Dulience, S. J., Joseph, S., Cooley, C., Tufte, T., Cox, K., Eaton, J., Delnatus, J. R., & Wolff, P. B. Fortified Snack Reduced Anemia in Rural School-Aged Children of Haiti: A Cluster-Randomized, Controlled Trial. PloS one, 2016; 11(12), e0168121. https://doi.org/10.1371/journal.pone.0168121

20. Wang, H., Zhao, Q., Boswell, M., & Rozelle, S. Can School Feeding Programs Reduce Malnutrition in Rural China? Journal of School Health, 2020; 90(1): 56–64. https://doi.org/10.1111/josh.12849

21. Abizari AR, Buxton C, Kwara L, Mensah-Homiah J, Armar-Klemesu M, Brouwer ID. School feeding contributes to micronutrient adequacy of Ghanaian schoolchildren. Br J Nutr. 2014 Sep 28;112(6):1019–33. doi: 10.1017/S0007114514001585. Epub 2014 Jul 3. PMID: 24990068.

22. Kwabla, M. P., Gyan, C., & Zotor, F. Nutritional status of in-school children and its associated factors in Denkyembour District, eastern region, Ghana: Comparing schools with feeding and non-school feeding policies. Nutrition Journal. 2018; 17(1). https://doi.org/10.1186/s12937-018-0321-6

23. Zenebe, M., Gebremedhin, S., Henry, C. J., & Regassa, N. School feeding program has resulted in improved dietary diversity, nutritional status and class attendance of school children. Italian Journal of Pediatrics. 2018; 44(1). https://doi.org/10.1186/s13052-018-0449-1

24. Jacoby E, Cueto S, Pollitt E. Benefits of a school breakfast among Andean children in Huaraz, Peru. Food and Nutrition Bulletin. 1996; 1: 54–64.

25. Agarwal DK, Agarwal KN, Upadhyay SK. Effect of mid-day meal programme on physical growth & mental function. Indian Journal of Medical Research. 1989; 90: 163–174.

